# Governance Framework and Public Private Partnership for Universal Health Coverage: Findings from India’s Federal Health Structure

**DOI:** 10.1101/2022.07.13.22277604

**Authors:** A. Venkat Raman, Chandrakant Lahariya

**Affiliations:** Faculty of Management Studies, University of Delhi, India; World Health Organization Country office for India, New Delhi and Foundation for People-centric Health systems, New Delhi, India

**Keywords:** Governance framework, Health Service Delivery, Indian States, Public Private Partnership, Universal Health Coverage

## Abstract

**Background:** The role of private health sector in advancing universal health coverage is being recognized globally. A number of public private partnership (PPP) strategies have been implemented across the states in India. States (provinces) are primarily responsible for health service delivery in India.

**Objectives:** To document various PPPs models in health sector across Indian states and to map the policy, legal and institutional eco-system governing such partnerships.

**Methods:** Desk review followed up with field visits and in-depth interviews. A total of 52 in-depth interviews were conducted from various levels of stakeholders.

**Results:** Nearly 250 PPP initiatives in health sector across all Indian states were identified and studied. Partnership with the private sector was predominantly in the areas of emergency transport, laboratory diagnosis, and in the delivery of selected primary care services. PPPs in health infrastructure (hospitals and medical colleges) and purchasing arrangements are rapidly emerging across most states. However, only few Indian states have health sector specific PPP policy or legal and/ or institutional framework governing PPPs and organizational units implementing partnership schemes. The capacity to conceive, design, implement, and manage PPPs in health sector was found either absent or insufficient in most states.

**Conclusion:** Effective partnerships with the private health sector for achieving country’s health goals requires a well enunciated policy and governance framework; detailed assessment of the private health sector market behavior; legal, regulatory, and oversight mechanisms; building organizational structures with capacities, and developing platforms for stakeholder dialogue. Findings from the Indian context could offer useful insights for other low- and middle-income countries aiming to advance towards achieving UHC.

## Introduction

Private health sector (PHS) delivers a large proportion of curative and diagnostic services in most parts of the world. PHS consists of both ‘for-profit’ and not -for-profit’ entities, ranging from Individual practitioners to quaternary care hospitals, delivering health care services. Household surveys across seventy developing countries indicate that 67% of diarrhoeal cases and 63% of fever cases among children are attended to by private caregivers.^1^ In the Association of South-East Asian Nations (ASEAN) region the private, for-profit sector accounts for more than 53% of the healthcare service delivery.^2^ In the 22 countries under Eastern Mediterranean Region (EMR) of the World Health Organization (WHO), the private sector accounts for 33% to 86% of outpatient services, with up to 81% of poorest quintile seeking care from the private sector. The proportion of private sector primary care facilities ranges from 20% to 90% in low income countries of the region.^3^ In the WHO regions, PHS accounts for almost 40% of all healthcare services in PAHO (Pan American region), AFRO (African region) WPRO (Western Pacific region), 57% in the SEARO (South-East-Asia Region) region, and 62% in the EMR region.^4^ In India, more than 75% of out-patient services and nearly 60% of hospitalization is sought from the private sector.^5^ The private sector has been growing steadily in many countries and has often been operating in parallel with the public sector health system.

In mixed healthcare system, which are reality in nearly all of low- and middle-income countries (LMICs), while accessing health services in private sector, people often have to pay from their ‘pockets’ (termed as Out of pocket expenditures or OOPE). Such payments are generally beyond the paying capacity of people, resulting in making them either poor or fall deeper into poverty if they were already poor. Alongside, given the relative strengths and limitations of both government and private sector, there is a growing recognition that healthcare needs of people could be met effectively, only if both sectors worked together^6-7^. Such arrangements are commonly referred to as Public Private Partnership (PPP). However, PPPs are professional contracts requiring skills and competencies as well as creating and sustaining enabling conditions and appropriate institutional framework - policy, legal and organizational arrangements- and capacities, to effectively govern such partnerships. Health service delivery and governance of the health system in India is decentralised and is primarily the responsibility of the states (provinces). Governance capacities and performance significantly vary across the states^8^. This study was conducted with an objective to summarise various PPP schemes and projects in health sector across the states in India; and to understand the eco-system i.e. policy, legal and institutional framework, that are critical for effectively governing the partnerships.

## Materials and Method

The study was carried out in two stages: a comprehensive desk followed by field visits for in-depth interviews of key stakeholders in select states. Desk review included compiling information on the policy, legal and institutional frameworks for PPP across the sectors, including the health sector, from the websites of relevant departments of the state governments (i.e. Planning, Finance, Infrastructure Development Authority, Health) across all the states and union territories. Information was compiled on whether that state / union territory has: i) State PPP policy; ii) PPP legislation; iii) PPP guidelines; iv) PPP cell; v) health sector specific PPP policy; and vi) health sector specific PPP cell. The desk review also included compiling information on public private partnership schemes and projects-in health service delivery, initiated by the states and union territories, from the respective websites of the department of health, medical education (in case of medical college-based PPPs), and of infrastructure development authority (in case of building health facilities). This was supplemented with reports of central and state ministries, development partners, key national level non-governmental organizations (NGOs), and industry associations. The review also included unpublished, draft versions of PPP policies, guidelines and reports. In addition, scientific literature on PPPs in LMIC context, were also reviewed. The information on PPP schemes/ projects was limited to the period from the years 2000 to 2016. Both ongoing and completed projects/ schemes were included in the listing. Some of the schemes involving the private sector in service delivery were excluded from the purview of this compilation and analysis. These are, country wide, centrally funded programs such as Rastriya Swathya Bima Yojana (RSBY), Employee State Insurance scheme (ESI), Central Government Health Scheme (CGHS), emergency ambulance services (EMR), and national disease control programs (e.g. TB, Malaria, HIV/AIDS). These were excluded as the state governments had limited role in the contract design or management of such PPPs. The analysis also excluded programs and schemes with NGOs in the areas of advocacy, community mobilization/outreach, information, education and communication (IEC), capacity building, monitoring and evaluation, grant-in-aid funding arrangements and projects under corporate social responsibility (CSR).

The desk review was followed by field visits to 13 states, selected on the basis of number of PPP schemes, to verify the PPP policy/ institutional ecosystem, as well as the status of the PPP schemes, and to collect additional details such as objectives of the partnership, scope of services, target beneficiaries, mutual obligations, etc. Interviews with 52 key stakeholders, such as senior government health officials and representatives of the private sector / NGO partner agencies, were conducted to gather feedback on the state’s experiences with PPP in health sector, including issues and challenges and lessons from the schemes. The interviews were conducted using an interview schedule, after obtaining formal permission from the state authorities. Information on each project was summarized in 200 words. The initial data collection (field visits) was undertaken in May-July 2017, and the data was further updated in March 2020. The data was analyzed using qualitative research methods, which has been described in an earlier publication^9^. The study was approved by the institutional ethics committee of faculty of management studies, University of Delhi, south campus. (Approval no. DU/IEC/ 2016/ MAY/101017)

## Results

More than 250 PPP projects and schemes in health sector were identified through desk review which was further verified through field visits to 13 states and through telephone calls with the officials of other states and union territories. All the states and union territories have piloted or implemented PPP schemes/ projects in health service delivery between the years 2000 to 2016.

In terms of policy (not restricted to health) 16 out of 35 states and union territories have a PPP policy either of their own or adopted the policy of the department of economic affairs (a central government agency). Only 10 states / union territories have a PPP legislation; and 9 states / UTs have PPP guidelines. Six states have both PPP policy and PPP legislation; PPP policy and guidelines are available in 6 states. Gujarat is the only state that has PPP policy, legislation, and guidelines. However, most states (26 states and union territories) have a PPP cell / unit for implementing the PPP projects across all sectors -not restricted to health. In many states, PPP cells were functional even without a PPP policy. Most of the PPP cells are independent agencies (11), followed by planning department (8) of the respective state or union territory governments. **(Table1)**.

**Table 1:** Overview of the Governance Eco-System for PPP/PSE in India.

| States | State PPP Policy | PPP Legislation | PPP Guidelines | PPP Cell | Health Sector PPP Policy | Health Sector PPP Cell |
| --- | --- | --- | --- | --- | --- | --- |
| **Andaman & Nicobar** | **D** |  |  |  |  | **TA** |
| Andhra Pradesh | D | √ |  | √ F |  | √ |
| **Arunachal Pradesh** | √ |  |  | √ P |  |  |
| Assam | √ |  | √ | √ P |  | √ |
| Bihar |  | √ | √ | √ A |  | √ |
| Chhattisgarh | √ |  | √ | √ F | √ |  |
| **Delhi** |  |  | √ |  |  |  |
| Goa | √ |  | √ | √ F |  |  |
| Gujarat | S | √ | √ | √ A |  |  |
| Haryana | √ |  | √ | √ A |  |  |
| Himachal Pradesh |  | √ |  | √ A |  |  |
| Jammu and Kashmir |  |  |  | √ F |  |  |
| Jharkhand |  |  |  | √ A | √ |  |
| Karnataka | √ | √ |  | √ A |  |  |
| Kerala | √ | √ |  | √ F |  |  |
| Madhya Pradesh | D |  | √ | √ F |  |  |
| Maharashtra |  |  |  | √ A |  | √ |
| Meghalaya |  |  |  | √ P |  |  |
| **Mizoram** | √ |  |  | √ P |  |  |
| Odisha | √ |  |  | √ P | √ | √ |
| Punjab |  | √ |  | √ A |  |  |
| Rajasthan | √ | √ |  | √ P | √ |  |
| Tamil Nadu |  | √ |  | √ F/A |  |  |
| Telangana |  |  |  | √ A |  |  |
| Uttar Pradesh |  |  | √ | √ P | √ | √ |
| Uttarakhand | √ | √ |  | √ P |  | √ |
| West Bengal | √ |  |  | √ F | √ |  |

D= Department of Economic Affairs (Federal Agency); S=Section Specific Policy; P = Planning Department; F = Finance Department; A = Independent Agency; TA = Transaction Advisors; No policy, legal and organizational framework in other provinces / union territories

Several states have adopted legal framework towards PPP in infrastructure development including health sector related infrastructure and investment opportunities in new health facilities, health spa, and health townships in some states. Many states indicated their preference to use Viability Gap Funding (VGF) as one of the preferred modes of promoting PPP in health sector infrastructure.

With regard to (health) sector specific policy and institutional framework, none of the states or union territories have a policy on private health sector (PHS), despite PHS accounting for majority of the service delivery in the state. Only 6 states have health sector specific PPP policy outlining strategies towards engaging the private sector. While health sector PPP policy is useful; but having a policy on PHS is more desirable in mixed health system like India, where it is critical to outline the role of PHS in achieving public health goals including universal health coverage. Such a policy would also spell out the government’s accountabilities with regards to licensing, regulation, accreditation, partnership and other oversight responsibilities. Lack of a PHS policy or PPP policy deprives the health sector the necessary legal, operational and regulatory framework in governing PPP projects/ schemes effectively.

Dedicated PPP cells were functional within the health departments of only 7 states and one union territory. These units are largely under-resourced (funds, staff), and poorly empowered (legal or managerial authority). The technical and managerial capacity of these PPP units to conceive, design, implement and monitor the partnership contracts is reportedly weak, leading to poor functioning of the partnership projects. This is compounded by poor regulatory compliance or enforcement and lack of legal framework for partnership governance.

Management contracts and service delivery PPPs are operational in every state of India. Many large states (more populated) have had the experience of PPPs in health infrastructure. In the recent past-before and during the Covid pandemic, health infrastructure PPPs have become a key focus area for many state governments in order to augment speciality wards, medical college cum hospitals, and diagnostic laboratories. Many states have state specific purchasing or government run social (health) insurance programs. Social (health) insurance and management contract PPPs co-exist in 11 states. In seven states, all three categories of PPPs are operational. Except medical college cum teaching hospitals, large scale hospital projects under PPP are not common in India. Management contract of government health facilities from primary health centres/ community health centres and district level hospitals have been in existence for more than two decades in some of the states. Similarly, many states have the experience of managing PPPs in the form of colocation of speciality wards, radio-dianoetic units, and laboratories; outsourcing of hospital support services (both clinical and non-clinical). Since the advent of Ayushman Bharat or Prime Minister’s Jan Arogya Yojana (PM-JAY), a country wide, centrally administered, social health insurance scheme, targeted to benefit low-income households, there has been a large scale purchasing of secondary and tertiary care services from the private health sector in many states (that adopted the scheme), since 2019. A summary of the scope and types of PPPs across the Indian states is given in **Table 2 and Box 1**, respectively.

**Table 2:** Scope of PPP Projects/ Schemes in Health Sector in India*.

| States | Infrastructure | Service delivery | Insurance / Financing |
| --- | --- | --- | --- |
| Andaman & Nicobar |  | √ |  |
| Andhra Pradesh | √ | √ | √ |
| Arunachal Pradesh |  | √ |  |
| Assam | √ | √ |  |
| Bihar | √ | √ |  |
| Chhattisgarh |  | √ | √ |
| Delhi |  | √ |  |
| Goa | √ | √ | √ |
| Gujarat | √ | √ |  |
| Haryana |  | √ |  |
| Himachal Pradesh |  | √ |  |
| Jammu and Kashmir |  | √ |  |
| Jharkhand | √ | √ |  |
| Karnataka | √ | √ | √ |
| Kerala |  | √ |  |
| Madhya Pradesh |  | √ | √ |
| Maharashtra | √ | √ | √ |
| Manipur |  | √ |  |
| Meghalaya | √ | √ |  |
| Mizoram |  | √ |  |
| Nagaland |  | √ |  |
| Odisha | √ | √ |  |
| Punjab | √ | √ | √ |
| Puducherry |  | √ |  |
| Rajasthan | √ | √ | √ |
| Tamil Nadu |  | √ | √ |
| Telangana |  | √ | √ |
| Tripura | √ | √ |  |
| Uttar Pradesh | √ | √ |  |
| Uttarakhand | √ | √ | √ |
| West Bengal | √ | √ |  |

The number of PPP projects /schemes in the states ranged from two to fifteen. The states with more than 10 PPP projects include Assam, Bihar, Odisha and West Bengal, Karnataka, and Rajasthan. Most common PPPs across the states include, contracting for radio-diagnostics and laboratory services, emergency response and referral transport, and provision of maternal and child health services, including birth deliveries.

In-depth interviews with key stakeholders revealed several operational level challenges while implementing and managing PPPs in health service delivery. Lack of in-house technical expertise (i.e. government health department) or a dedicated organizational unit (PPP cell) to conceive, design, implement and to supervise and monitor the PPPs is considered as the most significant challenge for PPPs in health sector by almost all government health officials. Officials handling the PPP related activities had either no prior experience or not given an opportunity to learn about PPPs through capacity building trainings. One of the senior official of the health department in a state stated, *“I am in the procurement department. One day my health secretary calls me and directs me prepare a full-fledged request for proposal (RFP) for a radio-diagnostic project in a district hospital, on PPP mode, within two days. Without any basic knowledge, I had to rely up on informal consultations with some ‘experts’ and internet sources to prepare the document which was modified by the health secretary. The iterations between me and my superiors went on till a time when the health secretary was satisfied. Then the proposal was put up for clearance from the legal and finance departments. That’s how I learnt about PPP. Now looking back, I wish I had some basic training on PPP and how to design professional contracts*.*”*

Without adequate expertise PPP projects are drafted by health department officials without due diligence on service specifications, quality indicators, performance metrics, oversight responsibilities, governance structure, financing mechanism, payment modalities, grievance and dispute settlement, etc. Contract management skills is another key area of concern. A district hospital health official stated, *“I have two PPP contracts running in my hospital. The project was conceived, designed and contracted by the state level officials. I played no role in the entire process neither there was any briefing or orientation on these contracts or how to manage them. How am I supposed to supervise and monitor them and verify their performance? I am also asked to verify their payment claims. How can I do that? The language in the contract is not easy for me to understand”*.

Although PPPs are being implemented for several years by the health department, most health officials admitted that they have no prior experience or exposure to PPPs. Some of the officials who worked on the PPPs were either promoted and transferred or retired. Capacity building training or knowledge transfer was virtually absent in most states. In some states, the service delivery contracts were designed by non-health departments (e.g. infrastructure development authority) or technical consultants of external agencies, who had little knowledge about contracting of clinical services. For example, in one state, management contract for a community health center was based on the rental value of the built-up space, rather than the scope of health services to be delivered from the facility. Expectedly after few months the contract culminated in to a legal dispute between the government and the private agency.

Both government officials and private sector representatives viewed PPPs as traditional procurement process of tendering. The slow and tedious process of tendering seem to have deterred many potential private partners. On few instances, the time taken for completing the tendering process was more than the duration of the contract itself. Tenders mostly focused on fulfilling eligibility conditions, rather than the any performance metrics. Stated one state level senior health official: *“The audit process primarily focuses on scrutinizing if the contract was issued to a private agency that meets all the eligibility requirements and to the L1 bidder (lowest priced bidder). Although we have started to use QCBS (Quality and Cost based screening) method in selecting a private agency but we have little understanding on performance-based contracting. As government officials we do not want any audit/ vigilance action for deviating from a conventional tendering process…*.*so what if contracts for procuring health services is completely different from procuring goods?”*

Poorly drafted contracts, not only affect the service delivery operations but also results in grievances and disputes. In some states, even complex service delivery contracts are signed on couple of pages; whereas in other states they are detailed at length. In both the cases grievances and disputes occur due to differing interpretations of the contract-the former being ambiguous and the latter being micro level details. Representatives of the private sector expressed concern over governments’ unwillingness to accord flexibility and autonomy in the overall management of service delivery operations. Contracts that define the governance structure for oversight role, are generally dysfunctional, as meetings are rarely held. Constant interference by health officials, often using threats and/ or strong-arm tactics under the guise of regulatory oversight, create conducive conditions for corruption to pay off the ‘regulatory’ harassment. Interference by local political activists for undue favors often leads to confrontation. Most contracts lack a defined mechanism for grievance redressal. Private sector representatives also highlighted instances where PPP contracts were either delayed or rolled back or not renewed or went under litigation due to opposition by interest groups and putting their investments at risk. Distrust on PPP in health sector is exacerbated by ideologically tinted discourse in the media, academics, and other pressure groups, resulting in an unfavorable environment for PPPs when in operation. Lack of stakeholder consultation, lack of information or dissemination on the features of PPP to the beneficiaries (at the community level), and poor accountability framework further compounds the distrust on PPPs in health sector. Lack of awareness among the beneficiaries often results in them being exploited by unscrupulous private partner agency staff to levy user charges on the patients who are not aware of their eligibility for free services, often resulting in confrontation. Importance of creating awareness in the community and wider publicity on the PPP scheme was emphasized repeatedly by the government officials.

The trust deficit is seriously eroded by payment delays-sometimes lasting for years or demand for ‘kick-backs’ for the release of over-due payments from the government. This concern is unanimously expressed by the private sector representatives. One of the not-for-profit private agency representatives stated *“We are an NGO. Our resources are limited. For nearly 14 months, we have not received payments for the services delivered. We have to pay for the salaries of the staff and for the materials and supplies. We have been ‘running from pillar to post’ without any remedy. We just want to exit from this contract after getting our payments settled. If we exit now, the payment may never be realized”*. Delay in payments is primarily due to lack of separate financial / budgetary allocation (called as ‘line item’) for PPP projects or schemes. As a result, government is forced to ‘make-do’ with ‘adjustments’ from other budgetary heads often leading to audit objections. Some states have made progress in streamlining the payment mechanism. In one of the outsourcing contracts in a large state, the payments are released automatically through electronic transfer, based on percentage of achievements of key performance indicators (KPIs). Deductions are automatically made in case of under achievement of KPIs. The monitoring system, which is the basis of release of payments, is automated using a robust IT-based platform linked to a server at the state health department.

One of the key characteristics of many projects PPPs across the states is that, the proposal for partnership was conceived and developed by individuals on both the sides, i.e., a bureaucrat or a minister from the government and founder of a senior leader from the private agency, and a contract was signed after mutual negotiations. Termed as ‘relational contracts’ such arrangements are highly vulnerable to fail or unsustainable, as the bureaucrats often get transferred and the new incumbent may have different priority or the relationship (personal rapport) between the individuals may not be the same. It also leads to either significant modification in the terms of contract or even termination creating uncertainty among the private partners, who become unwilling to enter in to future partnerships with the government. This raises a key governance issue, that is, a need for an explicit, sector specific, policy commitment to public-private partnership. A policy-based approach conveys political and administrative commitment, and to evince confidence among the serious private providers. Other key governance related issues expressed by stakeholders include:

i. Monitoring and ensuring that the eligible beneficiaries for free treatment, are given equitable quality care, and not asked for co-payments especially for high value (high cost) diagnostic services such as CT scan, MRI, and surgeries. There is also an accusation that private providers in hospital contracts prescribe unnecessary diagnostic tests and perform surgeries leading to inflated bills. There are also complaints about non-fulfilment of contract obligations especially related to the volume of services to be rendered to the ‘free’ patient category. There is always a dispute over the interpretation of ‘what is free and what is not’.
ii. Tariffs are fixed either on the basis of an archaic system followed by the state/ central government or based on lowest bid value. Tariffs for services such as lab investigations, fee for out-patient consultation, radio-diagnostics and surgical procedures are low and unviable for the private agency leading to compromise in the quality of services.
iii. Antipathy and animosity of the government staff towards the private agency is cited as another common concern, leading to poor coordination and lack of accountability of each other’s role. The government staff fears privatisation and job-loss or transfer. Private sector representatives also complained that wherever a co-location contract (i.e. managing part of a hospital clinical service unit), adequate infrastructure, utilities and amenities are not being provided causing severe inconvenience to the patients.

Stakeholders were also asked questions (through a brief questionnaire) with regard to their perception of PPPs in health sector. The questions included priority areas for PPPs, benefits and risks, and barriers to PPP. They were asked to rank 3 most critical barriers to PPP. Government health officials (n=41) indicated that, mutual suspicion and lack of trust between the public and the private sector, absence or lack of health sector specific PPP policy, and lack of capacity within the government to design, manage and monitor PPPs as the most important barriers. Private sector representatives (n=11) indicated two main barriers: lack of trust and mutual suspicion between the public and the private sector and lack of capacity within the government to design, manage and monitor PPPs. It is apparent that building trust and building capacities are the most important barriers for PPP in health sector.

## Discussion

There is a variable experience in all the states and union territories of India in implementing PPPs in health service delivery-some having had long experience, while others being at a nascent stage. However, lack of (health) sector specific policy, guidelines, and regulation on private partnership seem to be one of the main causes for ineffective implementation of the PPPs. A systematic review observed that, PPPs in health sector in India exist in a hostile eco-system and a deeply divided ideological discourse on the role of private health sector in health service delivery^6^. The heterogenous nature of private health sector and lack of information on them, poor licensing regime, weak regulatory enforcement, and lack of organizational capacity pose additional challenges in fostering sustainable collaboration with the private health sector. Lack of policy, legal and institutional framework as enabling conditions for PPPs in health sector, gives little confidence to the private sector to be involved in the PPP projects/ schemes. Without these foundations, PPPs may remain an ad-hoc solution to current health service delivery bottlenecks. These foundations are perhaps critical for effective governance of partnerships. The challenges identified in this study on PPPs in health sector are not unique to India and published literature across LMICs highlight similar concerns.^10-13^

During the past few years, both globally and in India, there has been a renewed interest to explore the policy option of engaging the private health sector (or PPPs) in health service delivery. Recognizing the critical role of the private health sector, the 63rd World Health Assembly (2010; Resolution #WHA 63.27)^14^ stated that *“…. (a) variety of private providers… play a significant and growing role in health-care delivery across the world… (however) governments… are faced with the challenge of constructive engagement with the complex range of health-care providers… in many countries effective engagement, oversight and regulation of private health-care providers may be constrained by imperfect intelligence, limited financial influence and weak institutional capacity…. building trust and constructive policy dialogue are vital for successful engagement, oversight and regulation”*. (Pg.59-61) The resolution urges member states to gather credible information, build institutional capacity and strong policy dialogue process for productive engagement with the private sector to ensure universal access to health-care services.

In the Indian context, the National Health Policy of India (2017) proposes the engagement of the private health sector in secondary & tertiary healthcare services^15^. In the last few years, the NITI Aayog (National Institute for Transforming India -a central government think tank), released model concession agreements (MCA) for engagement of private sector for delivery of select Non-Communicable Diseases (NCD) services^16^ as well as for establishment of Medical Colleges in remote districts using PPP option^17^. Clearly, the interest in PPP is on the rise in India.

However, engaging the private health sector is fraught with many challenges both in India as explained in the above paragraphs, as well as in most low- and middle-income countries. Asian Development Bank^18^ highlights some of the key challenges for PPPs in health sector, in the context of LMICs. Common challenges include (a) poor understanding of the concept of PPP; (b) weak institutional capacity of public sector agencies to engage in PPP; (c) PPPs being initially donor-driven and eventually losing momentum as the donors withdraw support; (d) political affinities and inability to sustain the PPP beyond the term of the ruling dispensation; (e) non-formal working arrangements between partners, which can result in limited support from one or both partners; (f) limited sustainability of resources; (g) lack of or weak monitoring; and (h) prevalence of moral hazards and political influences and practices. A report by KPMG^19^ found that PPPs have failed to achieve desired objectives due to selecting wrongs kinds of priorities (and projects) for PPP; poor definition of objectives; choosing wrong partners; making erroneous or overly restrictive assumptions and projections; inappropriate allocation of risks, and failing to generate sufficient competition. Many of these challenges are related to the governance aspects. Challenges that act as barriers for PPPs in LMICs, based on the review of scientific papers and reports from various countries, is summarised in **Box 2** ^10-13,20-28^.

Notwithstanding these challenges, evidence suggest a number of enabling conditions that could make PPPs more effective. Caribbean Development Bank^22^ proposed a few steps for making PPPs more effective. These are: (1) develop PPP policies, priorities, and establish the processes; (2) create enabling legal environment; (3) create institutional mechanism and build institutional capacity; (4) create organizational structure and develop human resource capacity and skills; and (5) allocate dedicated resources and create fiscal management and accounting frameworks. These steps have been proven to be effective in several countries across Europe and Australia in implementing PPPs. Considering the challenges in many LMIC countries are similar, these steps outlined above may be useful. In advancing towards UHC by engaging the private health sector in service delivery, the WHO (2020)^29^ has proposed six key governance behaviours critical to aligning private health sector with overall health systems goals. These are, i) build understanding; ii) deliver strategies; iii) enable stakeholders, iv) foster relations, v) align structure; and vi) nurture trust. The findings from this study in India, are similar to the challenges highlighted from other parts of the World. Based on the evidence from India and from other countries, the strategies proposed to improve enabling conditions for PPP in the low- and middle-income countries are listed in **Box 3**.

In 2020, during the response to the COVID-19 pandemic, the private sector played a key role in not only in the provision of clinical services (testing and treatment) but also other support services including vaccines, medical devices, and drugs and supplies. However, there had been instances when government had to use regulation overreach to ensure the services in private sector are affordable and available to all citizens.^30^ Although this is not a planned PPP, but underscores the potential contribution of the private sector in health service delivery if governed effectively.

As India aims to accelerate its progress towards UHC, it has to strengthen its governance capacity and tools to harness the private sector more effectively, as evident from this study.

## Conclusion

The importance and relevance of engaging private health sector for advancing universal health coverage is increasingly recognized. Strengthening policy, legal and institutional framework for governing the private health sector engagement is critical for successful and sustainable partnership. All states across India are engaged in one or more private sector partnership in health service delivery. This study highlights the need for developing appropriate policy, legal and institutional framework; organizational units with commensurate technical and managerial capacity within the health ministry to conceive, design and manage partnership contracts; developing enabling eco-system including stakeholders dialogue forums; and provide effective regulatory oversight including information on the private sector in order to govern partnerships. Such approach seems to be necessary across all mixed health systems in the low- and middle-income countries, where health sector governance in general is weak. In federalized health systems such as India, where states have limited capacity for PPP, the institutional mechanisms for regular capacity building need to be established.

## Data Availability

Yes. data will be made available on request.

## Acknowledgement

The authors would like to thank Dr Hilde de Graeve, Team Leader, health Systems for inputs on an earlier version of this manuscript.

## Annex

### Box 1

**Various types of PPP in Indian states**

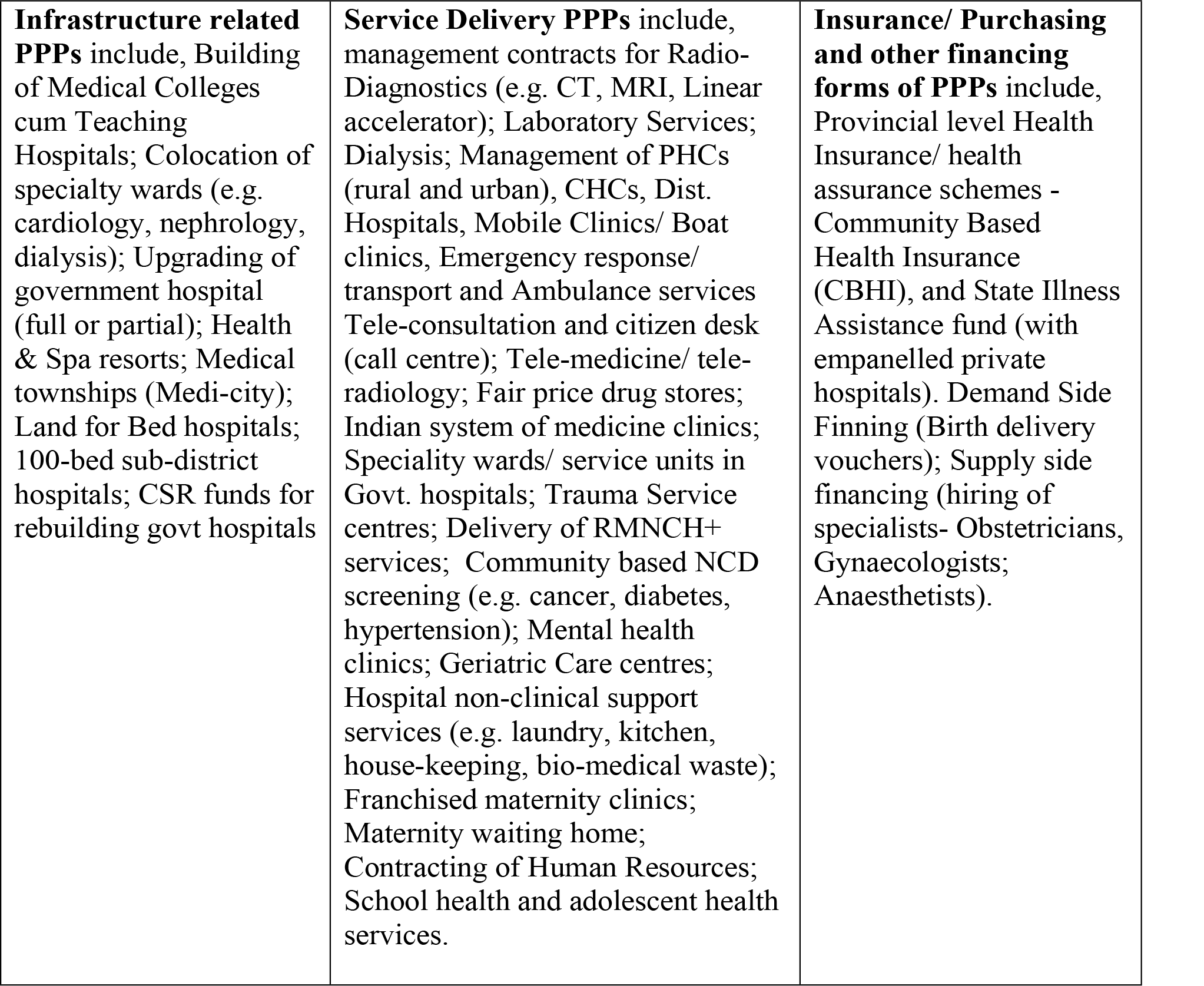

### Box 2

**Barriers in private sector engagement and PPP in low- and middle-income countries**

▪ Lack of explicit, long term political commitment and lack of enthusiasm from top bureaucracy towards private sector engagement in health sector;
▪ Inadequate information on the private health sector, its characteristics, standards, resources deployed, etc.
▪ Inadequate or absence of sector specific policy, legal, and regulatory framework and/or guidelines for engaging the private sector in health care;
▪ Weak or absence of organizational unit or technical and managerial capacity to conceptualise, design, implement, manage, and monitor projects or schemes related private sector engagement;
▪ Weak or inadequate rigour and organizational capacity to enforce provisions of licensing, regulation, physical standards, quality assurance and patient safety;
▪ Inadequate or even absence of dedicated budgetary provisions for private sector engagement projects and schemes;
▪ Lack of dialogue forums between the public sector and the private sector that perpetuates trust deficit and popular antagonism on the role of private providers in health sector.
▪ Tendency to use PPP or PSE as an ad-hoc solution to a current health sector problem;
▪ Delayed payments due to poor managerial capacity or lack of dedicated funds for the PPP project.
▪ Lack of grievance or dispute settlement system that creates ideal condition for corruption;
▪ Inadequate attention or due diligence to the project design, contract details leading to faulty projections or lack of flexibility (e.g. duration, cost, risks, performance indicators, tariff revision, replacement of equipment, etc.);
▪ Rolling out PPP project without piloting and learning before scaling up;
▪ Non-adherence to contract obligations by the public sector;
▪ Inadequate publicity and creation of awareness about the PPP project, services and the benefits to the target population
▪ Lack of evidence through comprehensive evaluation of PPP projects. This has led to a wide range opinions, perceptions and speculations about effectiveness, merits and demerits of PPP, its sustainability.

### Box 3

**Strategies suggested to improve and enable conditions for PPP/PSE**

▪ **Political Commitment and consensus amongst key stakeholders for Private Sector Engagement:** In most LMICs, health care (delivery) is traditionally seen as the responsibility of the government; and engaging the private sector would be criticised as an abrogation of government responsibility or even alleged privatisation, by stakeholders. Therefore, political and bureaucratic commitment towards engaging the private health sector is the first step towards institutionalising private sector engagement.
▪ **Assess the Private Health Sector**: Any policy on private health sector or private sector engagement, should begin with a careful review of their role in the country’s health system. Questions such as, (a) who the private health sector is; (b) what their characteristics are; (c) how and why they are relevant; (d) what their potential role could be; and (e) what kind of policy instrument is needed to engage them, etc, are critical for any strategy towards the private sector. Absence of such information could impede developing appropriate plans or strategies to engage them according to the contextual needs or where partnership with private sector may be more beneficial.
▪ **Adopt a Health Sector specific Private Sector Policy including PPP Policy:** A policy-based approach towards engaging the private sector, provides clarity on the overall objective and purpose. Governments may wish to formulate a broader ‘Policy towards Private Health Sector’ rather than just PPP policy for health sector to indicate its strategic priorities. Besides the policy, it is equally important to develop appropriate guidelines and provide legal sanctity to the policy.
▪ **Move from ‘ad-hoc’ to ‘institutionalized approach’ to PPP:** PPPs are legal and professional contracts and cannot be based on ‘relational contracts or contracts of good faith’. A policy on private health sector or PPP should be complemented with legal, regulatory and institutional framework with guidelines and governance structures.
▪ **Organizational Structure, Capacity and Resources:** Planning, designing and managing PPPs require a dedicated set of competent staff who can constantly interact with the private providers. If PPPs are to be managed well, it is important to create a separate unit or cell, with adequate staff and resources, with a mandate to implement private health sector policy or PPP policy. Attracting and retaining staff with certain degree of expertise in PPP in health sector is not easy for the government. Therefore, governments should organize capacity building programs and bring-in subject matter experts to ‘hand-hold’ the PPP cell staff in early stages of the functioning of the PPP cell or unit. Beside the organizational unit, government should allocate adequate budget for managing the PPP projects.
▪ **Regulatory Framework and Oversight:** The private sector cannot wish away the regulatory oversight by the government. Government also cannot fail in its obligations to enforce regulations or correct the private health sector behaviour. However, in many countries including India, the term regulation is perceived (by the private sector) as punitive, overbearing interreference by the government. This cognitive image of regulation should be redefined, and regulation needs to be portrayed as a harmonious functioning of the health system adhering to quality standards, rationality, equitable access, patient safety, accountability and efficiency in the interest of the country’s health system. Self-regulation precedes regulatory enforcement. Government and the private health sector should jointly develop regulatory guidelines and promote compliance-through both incentives and disincentives. PPPs could be used as incentives. But enforcement requires consistency and strong legal and institutional framework.
▪ **Build Dialogue Forums and ‘Community of Practice’ for knowledge sharing:** There is a trust deficit and the government officials, health staff, health activists, academics and policy analysts view the private sector with scepticism and its motives with suspicion. The private sector on the other hand is equally averse to any collaboration with the government due to overbearing or even hostile attitude and bureaucratic control. The trust deficit can be removed, only if the government and the private sector could interact with each other more frequently on matters relating to their expectations and concerns, in a candid manner. Such platforms for dialogue and consultation should be institutionalised.

## Notes

<u>Conflicts of interest:</u> There are no conflicts of interest related to this article.

### Clinical Trial

NA

### Funding Statement

The work was funded by the World Health organization India. However, the funder had no role in final content and opinion are that of the authors.

### Author Declarations

The study was approved by the institutional ethics committee of faculty of management studies, University of Delhi, south campus. (Approval no. DU/IEC/ 2016/ MAY/101017)

